# Transcriptome-Wide Association Analysis Identifies Novel Candidate Susceptibility Genes for Prostate-Specific Antigen Levels in Men Without Prostate Cancer

**DOI:** 10.1101/2023.05.04.23289526

**Authors:** Dorothy M. Chen, Ruocheng Dong, Linda Kachuri, Thomas Hoffmann, Yu Jiang, Sonja I. Berndt, John P. Shelley, Kerry R. Schaffer, Mitchell J. Machiela, Neal D. Freedman, Wen-Yi Huang, Shengchao A. Li, Hans Lilja, Stephen K. Van Den Eeden, Stephen Chanock, Christopher A. Haiman, David V. Conti, Robert J. Klein, Jonathan D. Mosley, John S. Witte, Rebecca E. Graff

## Abstract

Deciphering the genetic basis of prostate-specific antigen (PSA) levels may improve their utility to screen for prostate cancer (PCa). We thus conducted a transcriptome-wide association study (TWAS) of PSA levels using genome-wide summary statistics from 95,768 PCa-free men, the MetaXcan framework, and gene prediction models trained in Genotype-Tissue Expression (GTEx) project data. Tissue-specific analyses identified 41 statistically significant (p < 0.05/12,192 = 4.10e-6) associations in whole blood and 39 statistically significant (p < 0.05/13,844 = 3.61e-6) associations in prostate tissue, with 18 genes associated in both tissues. Cross-tissue analyses that combined associations across 45 tissues identified 155 genes that were statistically significantly (p < 0.05/22,249 = 2.25e-6) associated with PSA levels. Based on conditional analyses that assessed whether TWAS associations were attributable to a lead GWAS variant, we found 20 novel genes (11 single-tissue, 9 cross-tissue) that were associated with PSA levels in the TWAS. Of these novel genes, five showed evidence of colocalization (colocalization probability > 0.5): *EXOSC9, CCNA2, HIST1H2BN, RP11-182L21.6*, and *RP11-327J17.2*. Six of the 20 novel genes are not known to impact PCa risk. These findings yield new hypotheses for genetic factors underlying PSA levels that should be further explored toward improving our understanding of PSA biology.

## Introduction

Prostate-specific antigen (PSA) is a serine protease of the human tissue kallikrein-related (KLK) peptidase family. Serum levels are commonly used as a biomarker for detection, monitoring, and risk stratification of prostate cancer (PCa).^1–3^ The process by which a small portion of the highly abundant PSA in the prostate is released into the blood and elevated in men prostate cancer is incompletely understood, but disruption of the prostate gland architecture by neoplastic transformation has been speculated as a possible mechanism.^1, 3, 4^ PSA levels can additionally be influenced by age, race, body mass index (BMI), infection, prostate volume, benign prostate hyperplasia (BPH), and germline genetics.^5, 6^

PSA screening has been used for over 25 years for the detection of PCa, which is the second leading cause of cancer death among men in the United States.^1, 7–10^ However, low test specificity and discrimination have complicated the use and interpretation of PSA as a screening tool.^9, 11^ Long-term outcomes data from large population-based randomized PSA screening trials show that it significantly reduces death from prostate cancer but also results in considerable overdiagnosis of low risk disease.^12, 13^ Screening might be improved were it to account for variation in PSA levels that is attributable to germline genetics rather than PCa. Twin studies and genome-wide association studies (GWAS) have estimated that 30-45% of serum PSA variation is influenced by hereditary factors.^14, 15^ A recent GWAS meta-analysis from our group identified 128 independent variants associated with serum PSA levels, which explained approximately 9% of genetic variation in PSA.^16, 17^

Additional genetic variation in PSA levels may be determined by analyses of genes, as opposed to individual variants. Such work could lead to the identification of regions or biological pathways that affect PSA levels and provide clarity on mechanisms underlying constitutional increases in PSA levels in the absence of carcinogenesis. Transcriptome-wide association studies (TWAS) using expression quantitative trait loci (eQTL) allow for identifying genes whose cis-regulated expression is associated with complex polygenic traits.^18–21^ We thus performed a TWAS of PSA levels based on summary statistics from a GWAS meta-analysis of 95,768 men without PCa to identify genes associated with PSA levels and prioritize them for functional investigation.^15, 22^

## Methods

### Discovery Populations

A total of 95,768 PCa-free men from the following five study populations were included: UK Biobank, Kaiser Permanente’s Genetic Epidemiology Research on Adult Health and Aging, Prostate, Lung, Colorectal and Ovarian (PLCO) Cancer Screening Trial, Vanderbilt University Medical Center’s BioVU, and Malmö Diet and Cancer Study. The men included in these analyses were restricted to PCa-free individuals (i.e., without a PCa diagnosis or history of prostate resection, where information was available). Additionally, analyses were restricted to men with PSA values between 0.01 ng/mL and 10 ng/mL. Additional details about the study cohorts are described in detail in Kachuri et al. and Table S1. Median PSA levels were used for individuals with multiple PSA measurements available, with the exception of PLCO, which used PSA levels at the start of follow-up of the trial.

### Quality Control and GWAS Meta-analysis

Genotyping, imputation, study-specific quality control of genetic data, and the GWAS meta-analysis have been previously described for Kachuri et al.^17^ Briefly, ancestry and study-specific GWAS analyses used linear regression of log(PSA) on genetic variants, age, and the first 10 genetic ancestry principal components. After results across studies were meta-analyzed within ancestral groups, ancestry-specific summary statistics from individuals of European (n=85,824), African (n=3,509), East Asian (n=3,337), and Hispanic/Latino (n= 3,098) ancestry were meta-analyzed to generate multi-ancestry summary statistics.

### MetaXcan Transcriptome-wide Gene-based Analysis

We undertook a TWAS using the MetaXcan approach, which directly estimates Z-scores for associations between gene expression levels and PSA levels using meta-analyzed GWAS summary statistics with tissue-specific prediction models. Multivariate adaptive shrinkage (mashr) prediction models were trained on GTEx (version 8) eQTL data for whole blood and, separately, prostate tissue. The covariances and weights for models trained on individual tissue types were from the PredictDB Data Repository (https://predictdb.org/).^23^ We also undertook a cross-tissue analysis across 45 GTEx tissues that excluded tissues found primarily or exclusively in women to identify potential associations with genes that may have been missed in single-tissue analyses. Tissue types found primarily or exclusively in females (mammary breast, ovary, uterus, vagina) were excluded from the multiple tissue analysis. A Bonferroni correction was implemented based on the number of genes tested (whole blood: 0.05/12,192 =4.10e-6; prostate: 0.05/13,844 =3.61e-6; cross-tissue: 0.05/22,249 =2.25e-6).

### Conditional Analysis Ascertaining Independence of TWAS and Previous GWAS Findings

To identify the set of TWAS genes for PSA that were novel in comparison to previous GWAS results, we first limited consideration to significant TWAS genes that do not contain genome-wide significant (p < 5e-8) variants from the Kachuri et al GWAS (i.e., within exact gene boundaries).^17^ Second, for the remaining genes, we performed conditional analyses using COJO^24^ that simultaneously modeled eQTLs for a given TWAS-identified gene and GWAS results. For individual tissue analyses, eQTLs used in the prediction models of TWAS-significant genes were selected from the tissue-specific PredictDB TWAS eQTL weight files for each gene. For the cross-tissue analyses, the maximum noncollinear eQTLs (variant inflation factor < 8 within a window size of 50kb) were extracted from all 45 GTEx tissues (i.e., to remove variants in linkage disequilibrium). COJO was performed using LD reference panels from 10,000 European ancestry UK Biobank participants (as 89.6% of our population was European ancestry).^24^ GWAS summary statistics were then conditioned on the eQTLs for one gene at a time. If the set of genome-wide significant variants was reduced after conditioning on eQTLs used to predict gene expression, then the eQTLs and GWAS hits were assumed to be in LD and thus not independent of one another. (Methods that assess the conditional fit of eQTLs on GWAS hits are lacking.) However, if the set of genome-wide significant variants remained unchanged after conditional analyses, then we considered the TWAS gene to be independent of GWAS variants from Kachuri et al.^17^

### Colocalization Analyses

To further investigate whether genes and variants exhibited shared genetic signals, we used fastENLOC software (version 2) to undertake colocalization analyses of eQTLs for all genes with prediction models across TWAS analyses and all variants investigated in the Kachuri et al. GWAS.^17, 25, 26^ For these analyses, LD blocks were based on 1000 Genome Project European ancestry individuals.^27^ Z-scores were calculated by dividing the betas from GWAS summary statistics by their standard errors and converting them to posterior inclusion probabilities.^25^ Finally, the GWAS summary statistics were colocalized for 49 GTEx version 8 tissues using pre-computed GTEx multiple-tissue annotations. Signal-level results returned regional-level colocalization probabilities (RCP) between eQTL and GWAS signals, which sum up the variant-level colocalization probabilities of correlated variants within an LD block that harbors a single GWAS association signal. Gene-results were based on gene-level regional colocalization probabilities (GRCP), which represent the probability that a candidate gene contains at least one colocalized variant.^26^ RCP and GRCP values > 0.5 indicate strong evidence of shared genetic signals between eQTLs and GWAS variants. ^26^

### Pathway Enrichment Analysis

To explore the potential biological relevance of PSA-associated genes, we applied the Enrichr tool to all significant genes identified in the whole blood, prostate, and cross-tissue analyses to assess enrichment against three gene set libraries: Kyoto Encyclopedia of Genes and Genomes (KEGG) 2021 human; gene ontology (GO) biological process 2021; and GO molecular function 2021. Enrichment was assessed by multiplying the p-value from a Fisher’s exact test with the z-score of the deviation from the expected rank.^28, 29^ Pathways with a Benjamini-Hochberg-corrected p-value < 0.05 were considered significantly enriched.

## Results

Using MetaXcan and GWAS summary statistics based on 95,769 individuals (Table S1), TWAS analyses were conducted for whole blood, prostate, and cross-tissue matrices. The sample was primarily European (n=85,824), although individuals of African (n=3,509), East Asian (n=3,337), and Hispanic/Latino (n= 3,098) ancestry were also included in analyses. The median PSA value across all possible values across different cohorts was 2.35 ng/mL.

### Whole Blood TWAS

In the TWAS based on whole blood gene expression models, 41 out of 12,192 genes were associated with PSA levels at the Bonferroni corrected threshold (Figure 1, Table S2). Expression of 20 genes was positively associated with elevated PSA, and expression of 21 genes was inversely associated. Two of the significant genes are located at 19q13.33, which contains *KLK3*, the gene encoding serum PSA.^30^ While a prediction model for *KLK3* was not available in whole blood, a member of the same gene family at 19q13.33, *KLK2*, was the gene most strongly associated with PSA levels (p = 1.28e-62). Twenty-four of the 41 genes did not contain genome-wide significant variants and had not been annotated in previous GWAS. Among them, 7 genes were independent of previously published GWAS findings (Table 1).^17^ Of these, increased expression of 6 genes was associated with elevated PSA levels in PCa-free men: *GPBP1L1* (1p34.1, p = 2.21e-7); *TMEM69* (1p34.1, p = 2.21e-7); *UQCRH* (1p33, p = 4.76e-7); *ACTRT3* (3q26.2, p = 5.90e-7); *EXOSC9* (4q27, p = 1.80e-6); and *CCNA2* (4q27, p = 7.80e-7). Decreased expression of *ITH4* was associated with increased PSA levels (3p21.1, p = 3.65e-6).

**Figure 1.**
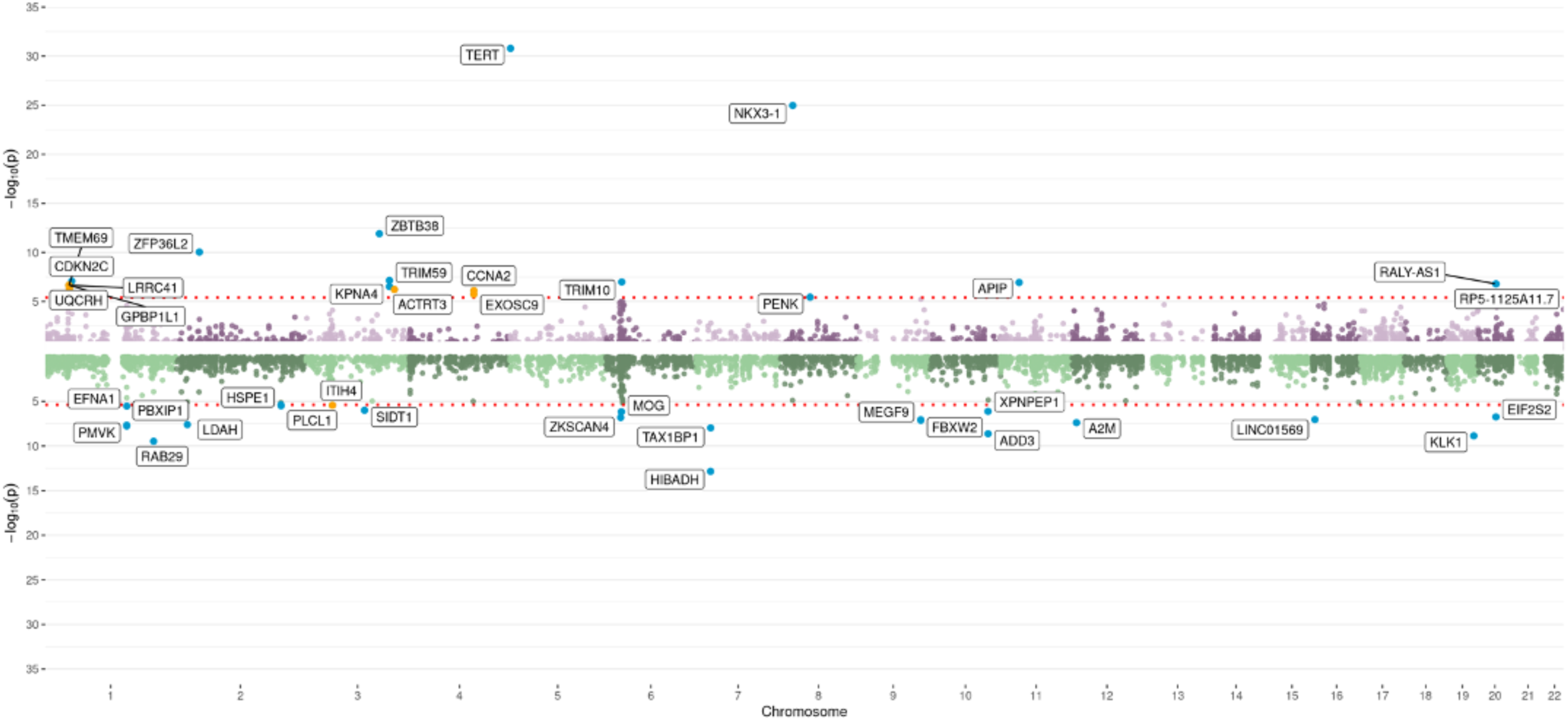
**TWAS results by chromosome**, as depicted by Miami plot for whole blood analyses. Each dot represents a gene, and the red dashed lines represent the significance thresholds after Bonferroni correction (whole blood: 4.10e-6, prostate: 3.61e-6, cross-tissue: 2.25e-6). For the Miami plots in figures 1 and 2, all statistically significant genes are annotated, and the genes independent of results from the largest prior GWAS are highlighted in yellow; the genes in the upper half of the plots were positively associated with PSA levels, and the genes in the lower half of the plots were inversely associated with PSA levels. For the Manhattan plot in figure 3, the 30 genes with the smallest association p-values are annotated and colored blue, and significant genes from the whole blood and prostate tissue analyses are annotated and colored yellow. Directions of association between genes and PSA levels are not available for cross-tissue analyses due to the joint tissue inference methodology used in S-MultiXcan.

**Table 1.**
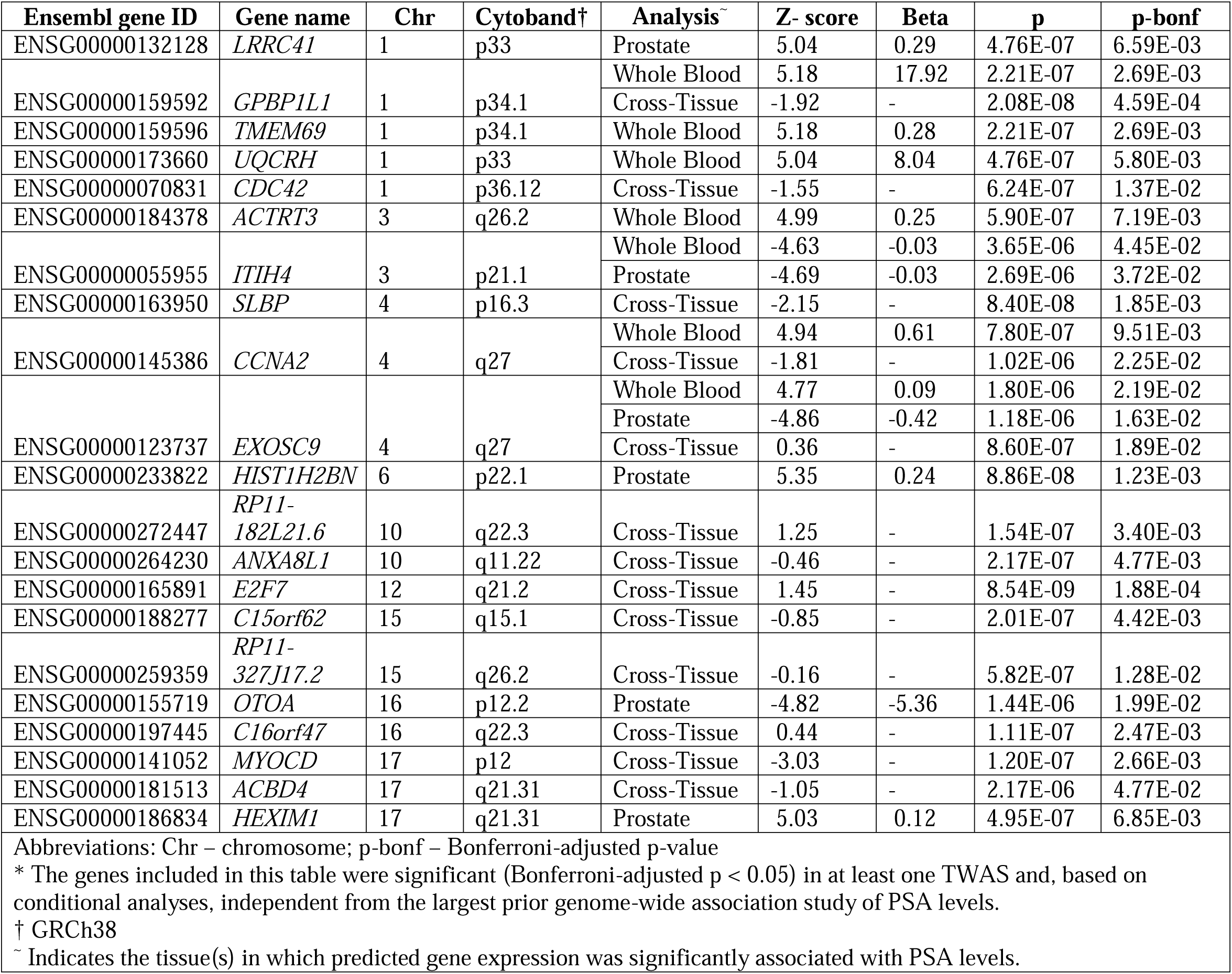
Novel genes associated with prostate-specific antigen (PSA) levels in transcriptome-wide association studies (TWAS) based on whole blood, prostate, and cross-tissue predicted gene expression*

### Prostate Tissue TWAS

In TWAS based on prostate tissue gene expression models, 39 out of 13,884 genes were statistically significantly associated with PSA levels (Figure 2, Table S3). Among them, increased expression of 18 and decreased expression of 21 was associated with elevated PSA levels. No prediction models were available for genes in the KLK family in prostate tissue. Instead, *MMP7* on 11q22 was most strongly associated with PSA levels (p= 2.78e-18). *MMP7* was among 18 PSA-associated TWAS genes containing known genome-wide significant variants. Of the remaining 21 significant genes, six were conditionally independent of previous GWAS results after conditioning on the eQTL variants used in gene prediction models (Table 1). Of these, increased expression of 3 genes was associated with elevated PSA levels: *LRRC41* (1p33, p = 4.76e-7), *HIST1H2BN* (6p22.1, p = 8.86e-8), and *HEXIM1* (17q21.31, p = 4.95e-7). Decreased expression of the remaining 3 genes was associated with increased PSA levels: *ITIH4* (3p21.1, p = 2.69e-6), *EXOSC9* (4q27 p = 1.18e-6), and *OTOA* (16p12.2, p = 1.44e-6).

**Figure 2:**
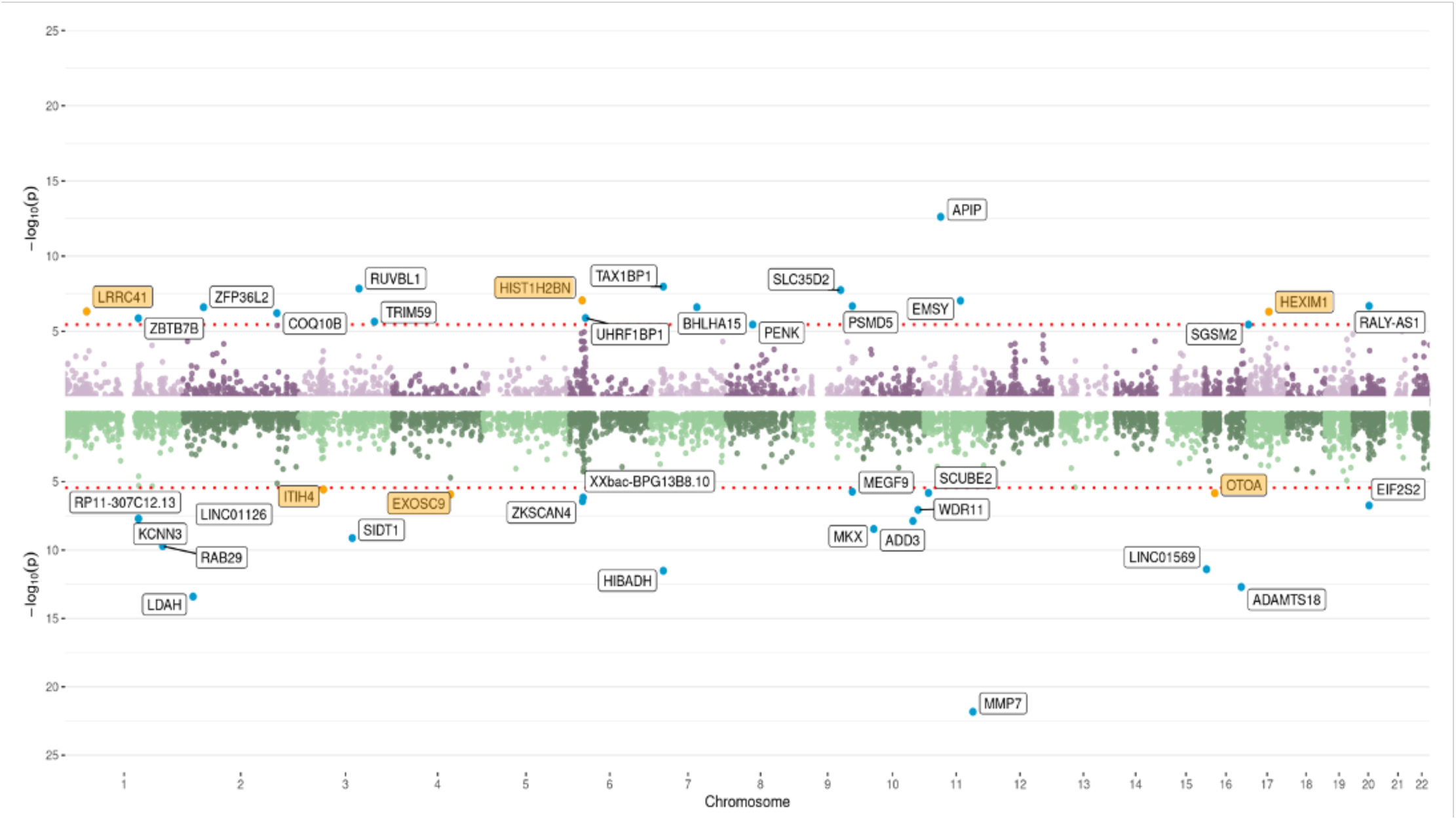
**TWAS results by chromosome**, as depicted by Miami plot for prostate tissue analyses. Each dot represents a gene, and the red dashed lines represent the significance thresholds after Bonferroni correction (whole blood: 4.10e-6, prostate: 3.61e-6, cross-tissue: 2.25e-6). For the Miami plots in figures 1 and 2, all statistically significant genes are annotated, and the genes independent of results from the largest prior GWAS are highlighted in yellow; the genes in the upper half of the plots were positively associated with PSA levels, and the genes in the lower half of the plots were inversely associated with PSA levels. For the Manhattan plot in figure 3, the 30 genes with the smallest association p-values are annotated and colored blue, and significant genes from the whole blood and prostate tissue analyses are annotated and colored yellow. Directions of association between genes and PSA levels are not available for cross-tissue analyses due to the joint tissue inference methodology used in S-MultiXcan.

### Cross-Tissue TWAS

Using S-MultiXcan to integrate signals from 45 tissues excluding tissues primarily or exclusively present in women, we identified 155 out of 22,249 genes whose predicted expression was associated with elevated PSA levels in cross-tissue matrices (Figure 3). Among the 155 statistically significant genes, 8 are located on 19q13.33 and part of the expanded human *KLK* gene family. We found 71 genes that do not contain genome-wide significant loci within their gene boundaries. Conditional analyses determined that 13 genes from this set were independent of known genome-wide significant variants: *CDC42* (1p36.12, p = 6.24e-07), *GPBP1L1* (1p34.1, p = 2.08e-08), *SLBP* (4p16.3, p = 8.40e-08), *EXOSC9* (4q27, p = 8.60e-07), *CCNA2* (4q27, p = 1.02e-06), *ANXA8L1* (10q11.22, p = 2.17e-07), *RP11-182L21.6* (10q22.3, p = 1.54e-07), *E2F7* (12q21.2, p = 8.54e-09), *C15orf62* (15q15.1, p = 2.01e-07), *RP11-327J17.2* (15q26.2, p = 5.82e-07), *C16orf47* (16q22.3, p = 1.11e-07), *MYOCD* (17p12, p=1.20E-07), and *ABCD4* (17q21.31, p = 2.17e-06) (Table 1).

**Figure 3:**
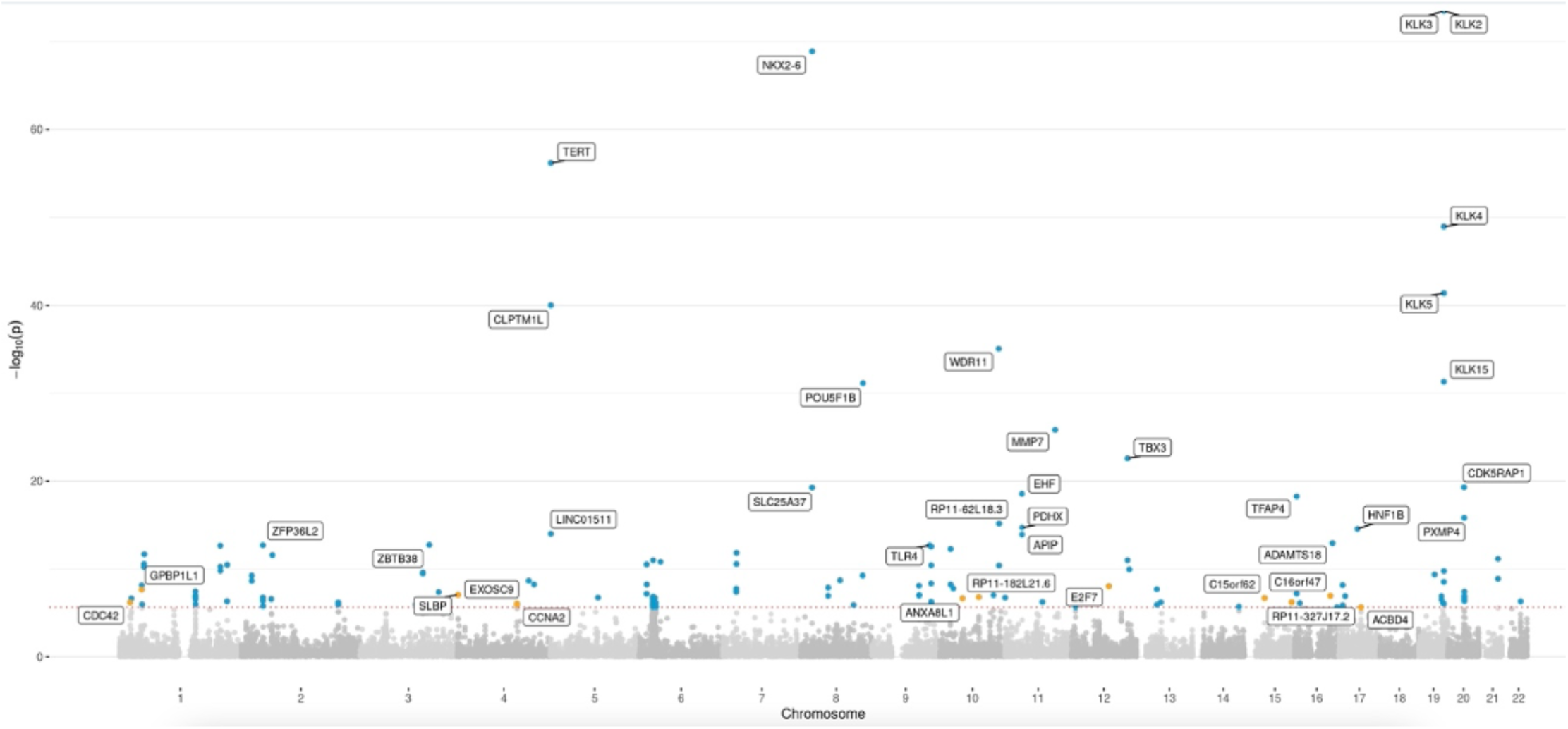
**TWAS results by chromosome**, as depicted by Manhattan plot for cross-tissue analyses. Each dot represents a gene, and the red dashed lines represent the significance thresholds after Bonferroni correction (whole blood: 4.10e-6, prostate: 3.61e-6, cross-tissue: 2.25e-6). For the Miami plots in figures 1 and 2, all statistically significant genes are annotated, and the genes independent of results from the largest prior GWAS are highlighted in yellow; the genes in the upper half of the plots were positively associated with PSA levels, and the genes in the lower half of the plots were inversely associated with PSA levels. For the Manhattan plot in figure 3, the 30 genes with the smallest association p-values are annotated and colored blue, and significant genes from the whole blood and prostate tissue analyses are annotated and colored yellow. Directions of association between genes and PSA levels are not available for cross-tissue analyses due to the joint tissue inference methodology used in S-MultiXcan.

### Significant Genes Overlapping Across TWAS

Out of the 41 genes that were significantly associated with PSA levels in the whole blood TWAS, 34 were also imputed in the prostate tissue TWAS. Thirty (30) out of 39 significantly associated genes from the prostate tissue TWAS were also evaluated in whole blood analyses. Of the 155 significantly associated genes detected in the cross-tissue analysis, 89 genes and 105 genes had transcriptome prediction models available for whole blood and prostate tissues, respectively.

Across all TWAS in whole blood, prostate tissue, and cross-tissue models, we identified 13 genes whose predicted expression was associated with elevated PSA levels: *RAB29* (1q32.1)*, LDAH* (2p24.1)*, ZFP36L2* (2p21)*, SIDT1* (3q13.2)*, TRIM59* (3q25.33)*, EXOSC9* (4q27)*, HIBADH* (7p15.2), *TAX1BP1* (7p15.2)*, MEGF9* (9q33.2)*, ADD3* (10q25.1)*, APIP* (11p13)*, LINC01569* (16p13.3), and *EIF2S2* (20q11.22) (Figure 4). Of these genes, only *EXOSC9* was conditionally independent after accounting for prior loci and demonstrated evidence of colocalization (described below). Five (5) associations were not statistically significant in the cross-tissue analysis, but had significant tissue-specific signals in whole blood and/or prostate. Seventeen significant genes were shared by only the whole blood and cross-tissue TWAS, six of which did not have prediction models in prostate tissue. Similarly, the prostate tissue and cross-tissue TWAS shared 14 significant genes; of these, seven genes did not have prediction models in whole blood.

**Figure 4.**
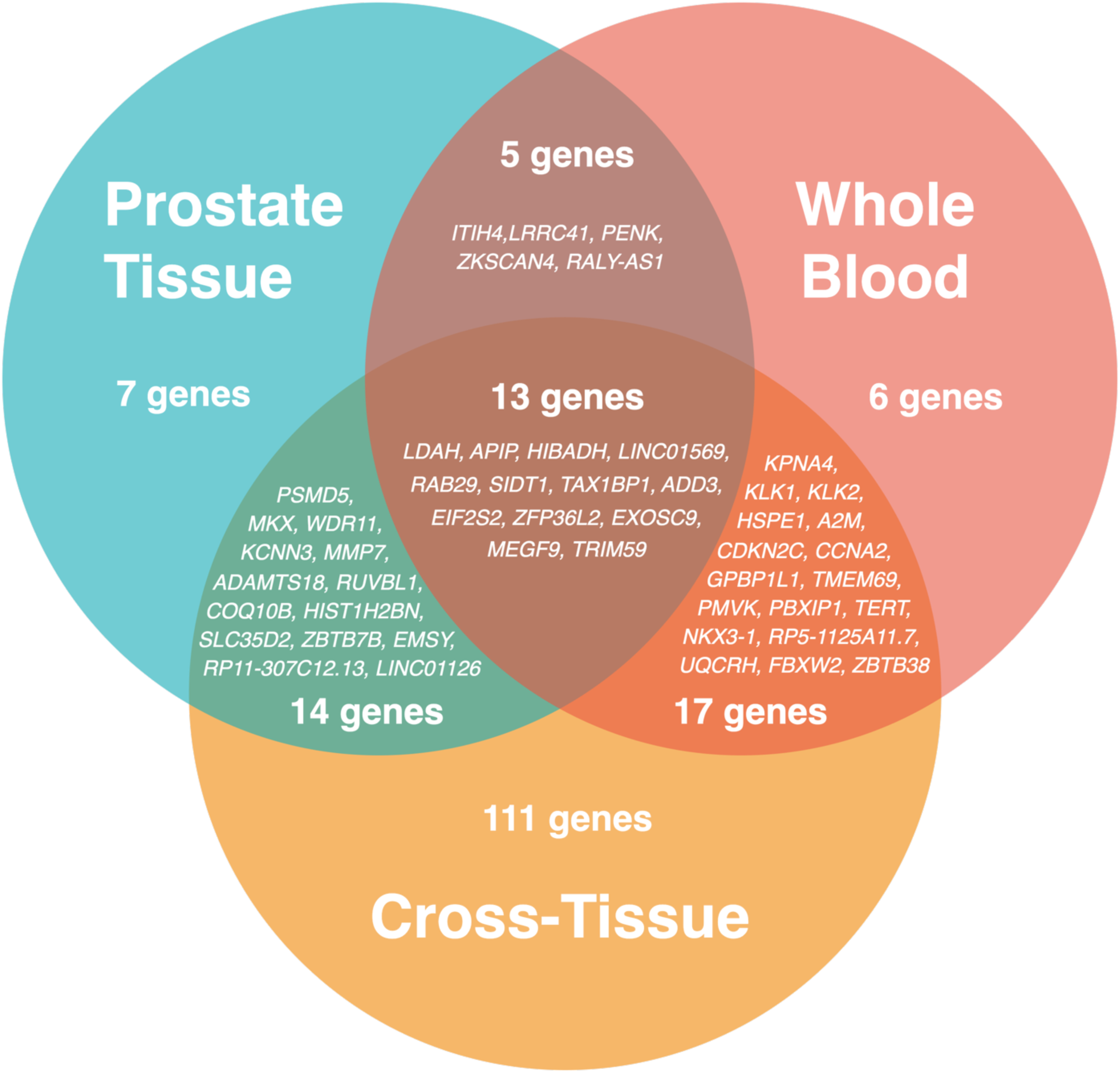
Statistically significant genes overlapping across TWAS analyses. Of the significant gene sets from whole blood (41 genes), prostate tissue (39 genes), and cross-tissue analyses (155 genes), 13 genes were significant across all three analyses, 36 genes were significant in exactly two analyses, and 124 genes were significant in a single analysis.

### Colocalization Analysis

Colocalization identified 372 signals with RCP > 0.5 at the variant level (Table S6) and 307 genes with GRCP > 0.5 at the gene level (Table S7). The top 10 genes with the largest GRCP were *OTX1* (2p15)*, MAFF* (22q13.1)*, FUT2* (19q13.33)*, EMSY* (11q13.5)*, IFT80* (3q25.33)*, EXOSC9* (4q27)*, SMC4* (3q25.33)*, RCAN3* (1p36.11)*, PBXIP1* (1q21.3), and *PMVK* (1q21.3). Of these genes, only *EXOSC9* was determined to be independent of previously discovered GWAS loci.^17^

Among 155 significant genes from the cross-tissue TWAS, locus-level results identified 50 genes with strong evidence of colocalization (Table 2). Variant-level results identified 66 regions in 51 genes (Table S5). When comparing the overlap between transcriptome-wide significant genes and results from colocalization analyses, we identified 5 novel genes (*EXOSC9, CCNA2, HIST1H2BN, RP11-182L21.6*, and *RP11-327J17.2*) with GRCP/RCP > 0.5. Two of these genes are long non-coding RNA while the remaining three are protein-coding genes.

**Table 2.**
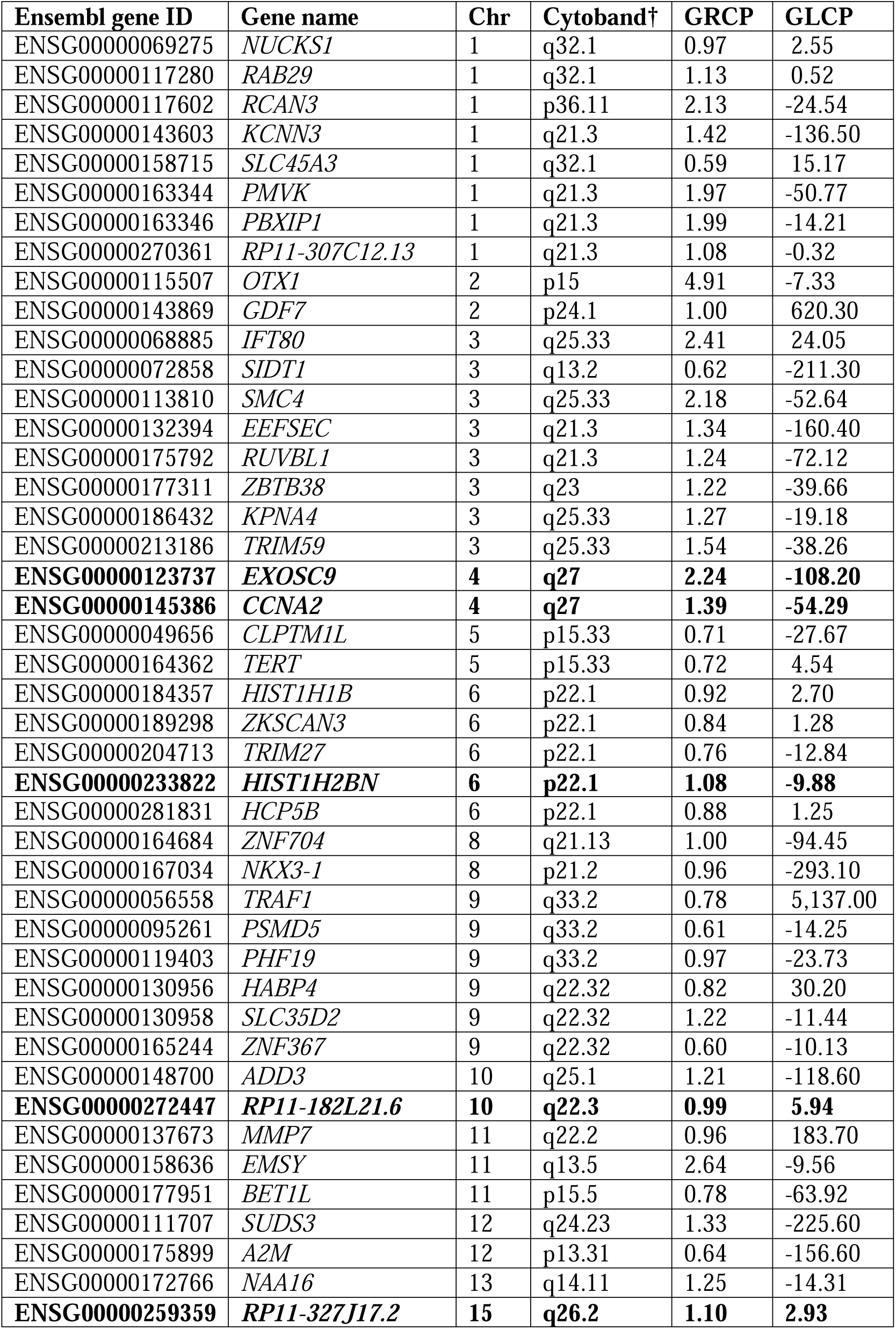
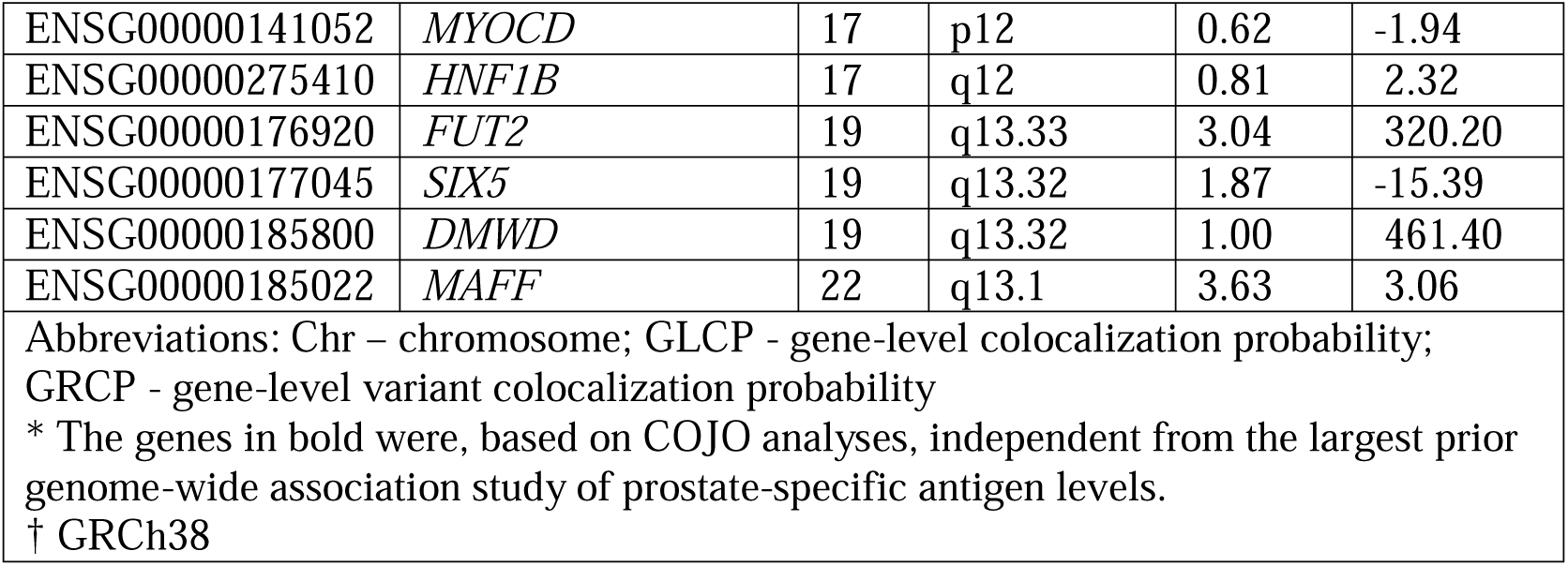
Gene level colocalization results for significant genes from the cross-tissue transcriptome-wide association study*

### Pathway Enrichment Analysis

TWAS genes were significantly enriched in 23 pathways underlying molecular activities and biological processes present in KEGG and GO gene set databases (Table 3, Figure 5). Enrichment analyses in the whole blood gene set identified the renin-angiotensin-aldosterone system in KEGG, which works to regulate arterial blood pressure. No pathways were significantly enriched by prostate and cross-tissue gene sets in the KEGG catalog.

**Figure 5.**
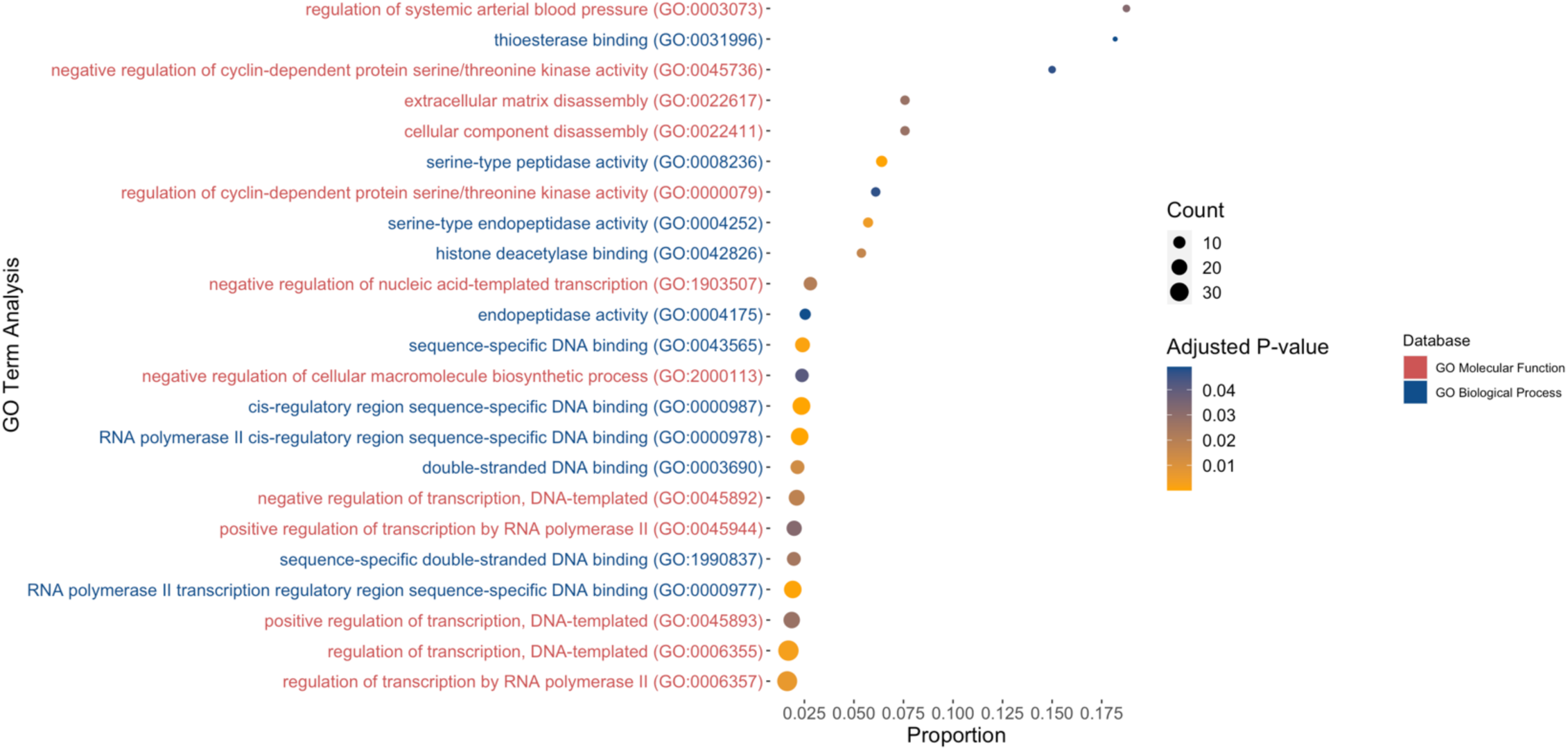
GO term enrichment results from the cross-tissue TWAS. The GO molecular function and biological process data repositories were queried for pathway enrichment analyses. The color gradient represents the magnitude of p-values, with darker colors corresponding to smaller values. The size of the circles represents the number of genes in the pathway.

**Table 3.**
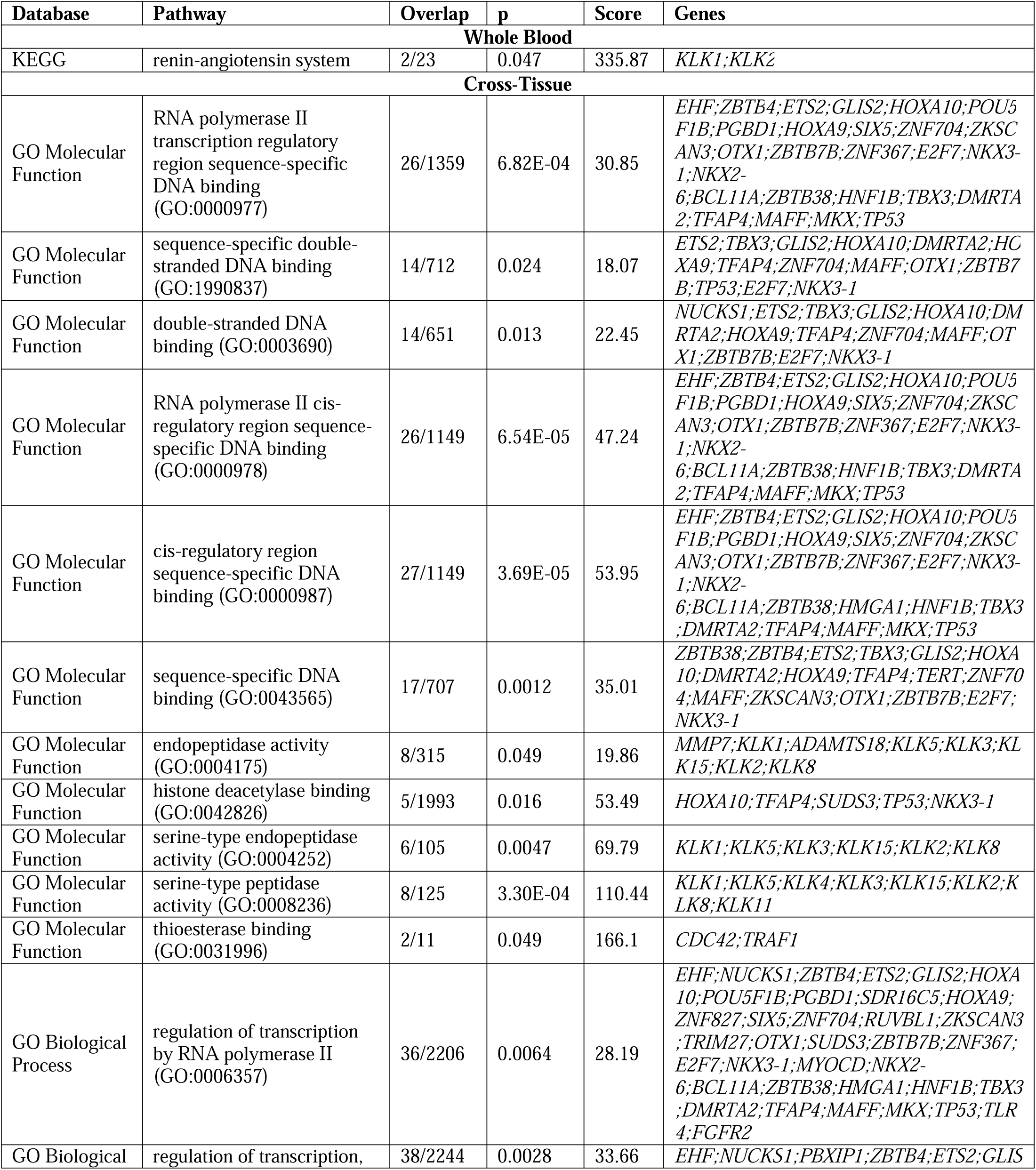
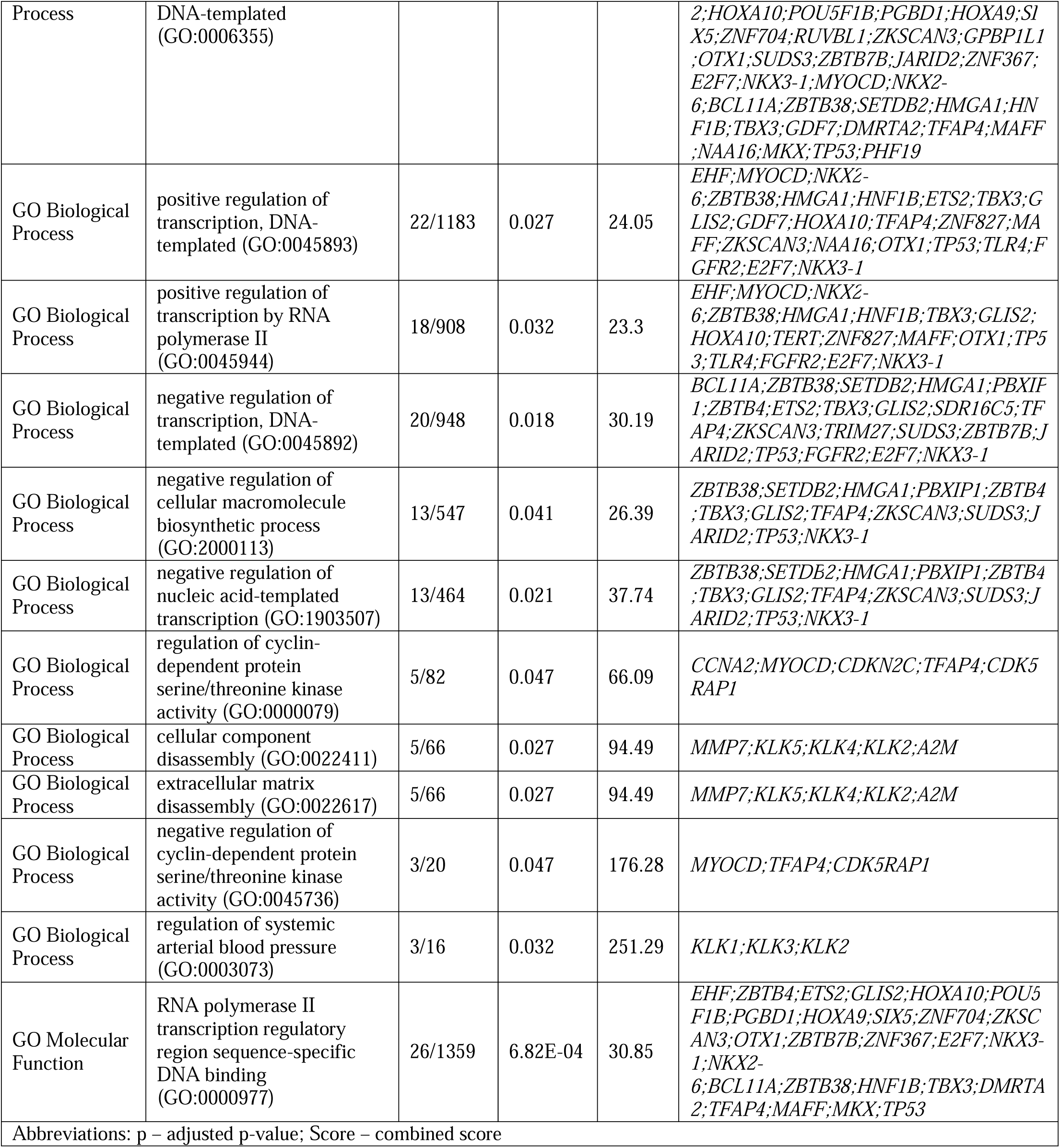
Results from pathway analyses based on all significant genes identified in the whole blood, prostate tissue, and cross-tissue transcriptome-wide association studies

Enrichment analyses within the GO biological process and molecular function gene set libraries identified 23 pathways that were significantly enriched in the cross-tissue gene set. No pathways were significantly enriched in GO for whole blood nor prostate tissue TWAS sets. Of note, the biological process underlying the regulation of cyclin-dependent protein serine/threonine kinase (CDK) activity in cell cycle progression is particularly relevant for the function of *CCNA2*. We also observed enrichment in 13 pathways involving both single and double stranded DNA binding mechanisms and regulation by transcription factors, including RNA polymerase II (pol II), a core component of the DNA transcription machinery. Moreover, two pathways underlying extracellular matrix and cellular component disassembly were enriched in the cross-tissue gene set. Analogous to the findings in KEGG for the whole blood TWAS, biological pathways underlying the regulation of systemic arterial blood pressure were also identified in the GO biological process catalog.

## Discussion

This TWAS of 95,768 PCa-free men identified 173 genes whose predicted expression levels were significantly associated with PSA. Conditional analyses identified 20 novel candidate susceptibility genes for PSA, and colocalization analyses highlighted five of them: *EXOSC9* and *CCNA2* at 4q27, *HIST1H2BN* at 6p22.1, *RP11-182L21.6* at 10q22.3, and *RP11-327J17.2* at 15q26.2. Pathway enrichment analyses across three gene ontology catalogs implicated regulatory pathways related to transcription, cell signaling, and disassembly of cellular and noncellular components such as the extracellular matrix.

The five colocalized genes introduce new hypotheses and insights regarding the genetic mechanisms regulating PSA production. Of particular interest are the signals at the 4q27 locus (*EXOSC9* and *CCNA2*), a genomic region that was not detected in previous GWAS of PSA.^16, 17^ Researchers have previously investigated an autoimmune related block on the 4q27 locus and did not find an association with PCa risk. ^32–35^ *EXOSC9*, which encodes a core protein involved in the RNA degradation machinery in humans, was identified across all three TWAS analyses. Currently, there is no literature supporting an association between *EXCOSC9* and PSA levels or PCa risk. Genetically predicted expression of *EXOSC9* in whole blood was positively associated with PSA levels, while the opposite direction of effect was observed for expression in prostate tissue. There exists limited tissue-specific knowledge of the physiological and pathological function of *EXOSC9* in non-cancer cell lines.^34, 35^ Literature documenting the relationship between *CCNA2* and PSA is also limited, though *CCNA2* was previously tested as a candidate autoantibody signature marker for distinguishing PCa from BPH in patients with elevated serum PSA;^36^ it was not determined to be a top antibody signature of any specific PCa targets. Gene co-expression network analyses have linked *CCNA2* to biochemical recurrence and survival in men with PCa.^37, 38^ Further exploration of *CCNA2* expression in individuals without PCa may unveil biological pathways that influence PSA levels.

Pathway enrichment results also highlighted a number of interesting pathways involving Pol II, which plays a critical role in the regulated synthesis of both protein-coding and noncoding messenger RNA in eukaryotic genomes.^39–41^ In addition to its numerous roles in DNA transcription, Pol II is known to associate with androgen receptor binding regions of the PSA enhancer and promoter to initiate androgen-dependent transcription.^42^ The two colocalized lncRNA genes, *RP11-182L21.6* and *RP11-327J17.2*, could feasibly regulate epigenetic modification through histone or DNA acetylation or methylation.^43^ That histone deacetylase binding was enriched in our pathway analyses suggests a possible mechanism through which lncRNAs could affect constitutive PSA levels. Aberrant expression of various lncRNA products has also been observed for various cancers, including PCa.^44^

Because PSA is used for both PCa detection and monitoring of PCa progression, it is difficult to disentangle the mechanisms underlying our observed associations. PSA-related screening bias may account for a portion of the observed relationship between gene expression and PSA levels. Genes may also exert pleiotropic effects on PCa and PSA through overlapping biological mechanisms.^1, 9^ Nevertheless, our restriction to PCa-free men for all analyses minimizes the potential for reverse causation and bolsters confidence that the observed genetic signals inform constitutive PSA levels. Fourteen of the 20 novel PSA-related genes identified from conditional analyses, spanning 12 genomic regions not implicated by prior PSA GWAS, have been associated with prostatic malignancies: *CDC42* (1p36.12), *GPBP1L1* and *TMEM69* (1p34.1), *LRRC41* (1p33), *ITIH4* (3p21.1), *SLBP* (4p16.3), *CCNA2* (4q27), *HIST1H2BN* (6p22.1), ANXA8L1 (10q11.2), *E2F7* (12q21.2), *C15orf62* (15q15.1), *OTOA* (16p12.2), *C16orf47* (16q22.3), and *HEXIM1*(17q21.31).^45–55^ Colocalized gene *HIST1H2BN*, which encodes a component of a core nucleosome histone, has been linked to PCa cell growth and epithelial-mesenchymal transition through upregulated NF-kB/Rel expression.^45^ Activation of the NF-kB pathway can induce activation of the PSA promoter-enhancer, even in the absence of androgens, and NF-kB can directly bind to the PSA enhancer in prostate cancer cell lines.^56^ To our knowledge, no experiments have been conducted to investigate the relationship between NF-kB and PSA in PCa-free populations.

Conditional analyses further identified 6 genes that have not been implicated in PCa susceptibility in gene function, experimental, or human population research: *UQCRH* (1p33)*, ACTRT3* (3q26.2)*, EXOSC9* (4q27)*, RP11-182L21.6* (10q22.3), *RP11-327J17.2* (15q26.2), and *ACBD4* (17q21.31)*. ACTRT3* is of particular interest, as it is critical in regulating sperm nucleus cytomorphology upstream of the processing of spermatid into mature motile sperm.^57^ Increased expression of *ACTRT3*, which forms a testis-specific profilin III-*ACTRT3* complex that facilitates male germ cell head cytomorphology and maintains sperm motility in animal models, was associated with elevated PSA levels in the TWAS based on whole blood.^57, 58^ Though there is no documented link between *ACTRT3* and PSA levels or PCa risk, the pronounced role of *ACTRT3* in mediating conformational changes in sperm nuclei suggests possible shared biological pathways between PSA and the production and processing of male germ cells.^5, 57^ Loci that map to the multigenic region that contains *ACTRT3* have been linked to melanoma, colorectal cancer, and lung cancer susceptibility.^59–61^ The functional role of *ACTRT3* is poorly characterized and warrants further analysis.

Our study had several limitations. First, nearly 90% of men in the study population were of primarily European ancestry, and 85% of the tissue samples used to train the TWAS models were derived from European populations.^62^ Therefore, our TWAS may have missed ancestry-specific signals, and our findings may not be generalizable to broader ancestral populations. To better characterize the genetic mechanisms underlying circulating PSA levels, it will be imperative for future analyses to expand to multi-ethnic populations; diversity in genetic studies of PSA levels is critical for equitably improving PSA screening.^63^ Second, our study assessed only cis-eQTLs, so any trans-eQTL effects are not incorporated. Third, colocalization analyses have a high type 2 error rate and may be underpowered to detect shared association signals.^26^ Fourth, although we restricted our analyses to men who had not been diagnosed with PCa, we cannot rule out the possibility of latent, undiagnosed disease or disease diagnosed at a later timepoint. However, the prevalence of undiagnosed PCa in our population was likely to be low on account of low prevalence at the time of the first PSA measurement or increased monitoring and surveillance in longitudinal cohorts.^7, 9^ Finally, many of the novel genes that we identified were clustered at multi-gene loci, in part due to co-regulation by a shared set of eQTLs. Sentinel genes at these loci should be interpreted with caution, as there may be correlated predicted expression.^64^

Our study also had several key strengths. The use of GWAS summary statistics from a cohort of 95,768 men provided us with high statistical power to quantify PSA-associated genes. In addition to conducting tissue-specific association analyses in tissues that are biologically meaningful for PSA, we integrated association signals across 45 GTEx tissues to improve power for genes with similar regulatory mechanisms across tissues. This method allowed us to capture PSA-related genes with expression patterns that were significant for specific tissues, as well as genes with expression that was similar across multiple tissues. Our study also used COJO analysis to find novel genes conditionally independent from known GWAS variants.

In summary, our TWAS identified gene expression profiles associated with PSA levels in men without PCa. These findings provide several novel hypotheses for genes that affect constitutive PSA. Further exploration of these results, including functional analyses of these genes in in-vivo settings, will augment our understanding of the genetic etiology of PSA variation. Transcriptomic analyses might also be vertically integrated with downstream -omic approaches to uncover complete mechanisms through which genetics influence circulating PSA levels. In addition, TWAS findings may be used to develop polygenic transcriptome risk scores^65^ for PSA levels, which could be leveraged for improving PSA as a tool for PCa screening.

## Data Availability

GWAS summary statistics used in this analysis are available for download from Kachuri et al are available from the following Zenodo repository: https://doi.org/10.5281/zenodo.7460134. Transcriptome prediction weights and models used for MetaXcan analyses are available from: https://predictdb.org.

https://predictdb.org

## Declaration of Interests

JSW is a non-employee, cofounder of Avail Bio. HL is named on a patent for assays to measure intact prostate-specific antigen and a patent for a statistical method to detect prostate cancer commercialized by OPKO Health (4KScore). HL receives royalties from sales of the assay and has stock in OPKO Health. HL serves on the Scientific Advisory Board for Fujirebio Diagnostics Inc and owns stock in Diaprost AB and Acousort AB.

## Funding sources

The Precision PSA study is supported by funding from the National Institutes of Health (NIH) and National Cancer Institute (NCI) under award number R01CA241410 (PI: JSW). REG is supported by a Prostate Cancer Foundation Young Investigator Award. LK is supported by funding from National Cancer Institute (R00CA246076). JPS is supported by funding from the National Institute of General Medical Sciences (T32GM007347). HL is supported in part by funding from NIH/NCI (P30-CA008748, U01-CA199338, R01CA244948) and the Swedish Cancer Society (Cancerfonden 20 1354 PjF). RK is supported by funding from the NIH (R01 CA244948 and R01 CA175491).

The content is solely the responsibility of the authors and does not necessarily represent the official views of the NIH.

## Notes

### Author Declarations

The Institutional Review Board of University of California, San Francisco gave ethnical approval for this work.

